# Prevalence, Predictors, and Persistence of Inadequate Spectacle Correction in Vietnamese Schoolchildren

**DOI:** 10.64898/2026.09.08.26362491

**Authors:** Nhan Thi Ho, Khoi Pham Minh Tran

**Affiliations:** Research Management Department, Vinmec International Hospital, Hanoi, Vietnam; Pediatric Department, Vinmec Times City, Vinmec International Hospital, Hanoi, Vietnam; College of Health Sciences, VinUniversity, Hanoi, Vietnam

**Keywords:** refractive error, spectacle, correction, adequate, inadequate, Vietnam, children, school health

## Abstract

**Background:** Inadequate spectacle correction limits the benefit of school vision screening, yet most evidence addresses only whether a child owns glasses rather than whether those glasses work. This study examined the prevalence, predictors, and persistence of inadequate spectacle correction among schoolchildren in three Vietnamese cities.

**Methods:** We analyzed annual vision screening records of students from a private school network in Ha Noi, Ho Chi Minh City, and Hai Phong, 2021 to 2024. Inadequate correction was defined as visual acuity below the age-appropriate threshold despite current glasses. A population-averaged modified-Poisson regression with robust standard errors, clustered by child, estimated adjusted prevalence ratios (aPR) for city, age, sex, and calendar year. Children linked across consecutive years allowed persistence analysis.

**Results:** Among 39,697 spectacle-wearing examinations, 30,936 (77.9%) were inadequately corrected, ranging from 64.8% (95%CI= 63.9, 65.8) in Ho Chi Minh City to 96.7% (96.0, 97.3) in Hai Phong. After adjustment, prevalence was lower in Ho Chi Minh City (aPR= 0.76; 95%CI= 0.75, 0.78; p<0.001) and higher in Hai Phong (aPR= 1.25; 95%CI= 1.23, 1.26; p<0.001) relative to Hanoi, and fell with age (aPR= 0.97 per year; 95%CI= 0.96, 0.97; p<0.001). Among 9,285 linked year pairs from 8,341 children, 85.3% (84.4, 86.1) of initially inadequately corrected children remained so the following year.

**Conclusions:** Most spectacle-wearing children in this Vietnamese cohort were inadequately corrected, with wide geographic variation and high persistence. School vision programs need mechanisms to verify correction adequacy after glasses are dispensed, not only to detect refractive error.

## Introduction

Uncorrected refractive error remains the leading cause of vision impairment in children worldwide, affecting an estimated 12 to 13 million children aged 5 to 15 years ^1–3^. Most of this burden is correctable with a simple pair of glasses, yet global consensus estimates suggest that roughly 12 million of the 19 million children currently living with vision impairment owe that impairment to refractive error alone ^2,4^. Spectacle correction is therefore one of the most cost effective interventions available in child health, and trial evidence shows its impact on learning outcomes can exceed that of other school health programs ^3^.

The burden of refractive error is not distributed evenly across the world. In East and Southeast Asia, myopia prevalence has climbed to 80 to 90% among young adults, with 10 to 20% progressing to high myopia beyond negative 6 diopters ^5,6^. This shift is driven largely by intensive academic schedules and limited time outdoors, a relationship strong enough that randomized trials of increased outdoor time have slowed the epidemic at a system wide scale ^5^. As myopia has become more common, anisometropia has followed a similar path, shifting from a condition traditionally associated with hyperopia to one increasingly tied to myopic refractive error, with downstream effects on stereopsis, binocularity, and rates of amblyopia ^7^. Regional data from India show a somewhat different pattern, with an overall refractive error prevalence of 8.0 per 100 children in population based studies, rising to 10.8 per 100 in school based surveys, and myopia concentrated in urban areas ^8^.

Detecting refractive error is only the first step. A growing body of evidence shows that spectacle correction itself improves educational and psychological well being, with systematic reviews reporting strong or very strong evidence for gains in reading fluency, academic performance, and quality of life once children are properly corrected ^3,9^. The inverse is equally well established. Children with greater severity of uncorrected refractive error and poorer visual acuity are more likely to actually wear the glasses they are given, while children with milder impairment often do not perceive a need for correction at all ^2,10^. This creates a paradox familiar to school health programs everywhere. Screening finds the children who need glasses, but simply handing out a prescription does not guarantee the child ends up seeing well. Reported barriers span cost, the quality of dispensed lenses, mistaken beliefs that glasses can harm a child’s eyes, and low perceived need among children with only mild refractive error ^1,10^. Despite decades of school based screening programs across low and middle income countries, most published evaluations still stop at whether a child received a pair of glasses rather than whether that correction was actually adequate ^3,10^.

Vietnam sits within the epicenter of the regional myopia epidemic, yet remains one of the least studied countries in this literature. A recent systematic review of refractive error and educational outcomes identified only a single Vietnamese study among twenty five eligible studies worldwide, leaving the country’s largest cities essentially undocumented with respect to spectacle correction adequacy ^11^.

To address this gap, our study performed retrospective analysis of multiple years of annual school based health check data which included vision screening of students attending a private school system from three major Vietnamese cities (Ha Noi, Ho Chi Minh City, Hai Phong). This large scale vision screening data offers a good opportunity to estimate the prevalence and correlates of inadequate spectacle correction among schoolchildren who wear glasses, and evaluates whether inadequate correction resolves or persists across consecutive annual visits.

## Methods

### Study design and population

We conducted an observational study using annual school health check data of students attending a private school system three major cities in Vietnam (Ha Noi, Ho Chi Minh City, and Hai Phong) from 2021 through 2024. The health check included standardized vision screening and assessment of refractive error and spectacle use. The primary analysis described inadequate spectacle correction among children wearing spectacles. A secondary longitudinal analysis examined changes in correction status between consecutive annual examinations. The study was reported in accordance with the Strengthening the Reporting of Observational Studies in Epidemiology recommendations ^12^. The study was approved by Vinmec Ethical Committee (approval number 0231/2024/CN/HDDD VMEC). Informed consent was waived for analysis of de-identified data.

Children aged 3 to 18 years with recorded examination year, examination site, and sex were eligible. School level was classified as kindergarten, primary school, lower secondary school, or upper secondary school.

### Definition of spectacle use and inadequate correction

The primary analytic population comprised children identified as currently wearing spectacles. Spectacle status was recorded during the vision screening examination. Inadequate spectacle correction indicated that a child wearing spectacles did not achieve adequate visual acuity with the current correction and vice versa.

This definition was chosen to describe the effectiveness of the child’s current spectacle correction at the time of screening. It does not by itself establish the optical cause of reduced corrected vision, such as an outdated prescription, an incorrect prescription, poor spectacle fit, non-compliance with spectacle use, amblyopia, or ocular disease. We therefore use the term inadequate spectacle correction rather than assuming that every case represents a clinically confirmed prescribing error. This distinction is important because effective refractive error services require both access to spectacles and appropriate quality of refractive correction ^13^.

### Statistical analysis

We first described the characteristics of spectacle-wearing children overall and according to correction status. The prevalence of inadequate spectacle correction was estimated overall and by examination year, city, sex, school level, and age group, with 95% Wilson confidence intervals.

Repeated observations from the same child were addressed using generalized estimating equations (GEE) with a log link and Poisson variance, robust sandwich standard errors, and an exchangeable working correlation structure. This approach provides population-averaged prevalence ratios while accounting for within-child correlation across annual examinations ^14,15^. The model included calendar year, continuous age, sex, and city. Adjusted prevalence ratios with 95% confidence intervals (95%CI) were reported. The modified Poisson approach was selected because prevalence ratios are more directly interpretable than odds ratios when the outcome is common ^15^.

Children with at least two valid examinations were eligible for longitudinal assessment. Same-year duplicate records with conflicting correction status were excluded from linkage. We restricted the primary transition analysis to consecutive annual examinations and classified each pair as adequate to adequate, adequate to inadequate, inadequate to adequate, or inadequate to inadequate. Row percentages were used to estimate the probability of each correction status at the subsequent annual examination. Among children who were inadequately corrected at the first examination, a separate GEE model was used to examine factors associated with persistence of inadequate correction at the subsequent examination.

Sensitivity analyses evaluated a broader refractive error definition and a restriction to children aged at least 6 years. Analyses were performed in R. All statistical tests were two-sided and a P value below 0.05 was considered statistically significant.

## Results

Between 2021 and 2024, 39,697 spectacle-wearing examinations were recorded across the three cities, representing children whose caregivers had already sought correction for a refractive error. **Table 1** summarizes the characteristics of this cohort. The children were evenly split by sex, with a mean age of 12.2 years. Most examinations came from Hanoi (68%), with smaller contributions from Ho Chi Minh City (24%) and Hai Phong (8.3%). Children with adequate correction were on average about a year older than those who were inadequately corrected (mean age 13.1 years versus 11.9 years), and adequately corrected children were also somewhat more concentrated in the upper secondary age range.

**Table 1.** Characteristics of spectacle-wearing children by correction adequacy, 2021 to 2024.

| Characteristic | Overall<br>N = 39,697<sup>#</sup> | Adequately corrected<br>N = 8,761<sup>#</sup> | Inadequately corrected<br>N = 30,936<sup>#</sup> |
| --- | --- | --- | --- |
| Sex |  |  |  |
| Male | 19,247 (48%) | 4,412 (50%) | 14,835 (48%) |
| Female | 20,450 (52%) | 4,349 (50%) | 16,101 (52%) |
| Age, years | 12.19 (2.87) | 13.11 (2.85) | 11.93 (2.82) |
| Age group |  |  |  |
| 3 to 5 years | 84 (0.2%) | 67 (0.8%) | 17 (<0.1%) |
| 6 to 11 years | 16,608 (42%) | 2,386 (27%) | 14,222 (46%) |
| 12 to 14 years | 13,179 (33%) | 3,095 (35%) | 10,084 (33%) |
| 15 to 18 years | 9,826 (25%) | 3,213 (37%) | 6,613 (21%) |
| School level |  |  |  |
| Kindergarten | 84 (0.2%) | 67 (0.8%) | 17 (<0.1%) |
| Primary | 11,933 (30%) | 1,650 (19%) | 10,283 (33%) |
| Lower secondary | 17,854 (45%) | 3,831 (44%) | 14,023 (45%) |
| Upper secondary | 9,826 (25%) | 3,213 (37%) | 6,613 (21%) |
| City |  |  |  |
| Hanoi | 27,055 (68%) | 5,371 (61%) | 21,684 (70%) |
| Ho Chi Minh City | 9,329 (24%) | 3,281 (37%) | 6,048 (20%) |
| Hai Phong | 3,313 (8.3%) | 109 (1.2%) | 3,204 (10%) |
| Year of examination |  |  |  |
| 2021 | 2,290 (5.8%) | 495 (5.7%) | 1,795 (5.8%) |
| 2022 | 14,647 (37%) | 2,833 (32%) | 11,814 (38%) |
| 2023 | 10,894 (27%) | 2,708 (31%) | 8,186 (26%) |
| 2024 | 11,866 (30%) | 2,725 (31%) | 9,141 (30%) |

Values are n (percent) for categorical variables and mean (SD) for age. Percentages within each column may not total 100 due to rounding.

Only a minority of children who wore glasses actually achieved an adequate visual outcome. Across all examinations, 30,936 of 39,697 spectacle wearers, or 77.9%, were inadequately corrected. This proportion varied sharply by city (**Table 2**, **Figure 1**). Hai Phong had the highest burden, with inadequate correction affecting 96.7% of spectacle wearers (95% CI 96.0 to 97.3). Hanoi followed at 80.1% (79.7 to 80.6), while Ho Chi Minh City had the lowest prevalence at 64.8% (63.9 to 65.8). As shown in **Figure 1**, this city ranking was stable across all four years of the study, with Ho Chi Minh City consistently below Hanoi and Hanoi consistently below Hai Phong.

**Table 2.** Stratified prevalence of inadequate spectacle correction.

| Stratifier | Stratum | n wearing glasses | n inadequate correction | Inadequate correction, % (95% CI) |
| --- | --- | --- | --- | --- |
| City | Hanoi | 27,055 | 21,684 | 80.1 (79.7 to 80.6) |
|  | Ho Chi Minh City | 9,329 | 6,048 | 64.8 (63.9 to 65.8) |
|  | Hai Phong | 3,313 | 3,204 | 96.7 (96.0 to 97.3) |
| Sex | Male | 19,247 | 14,835 | 77.1 (76.5 to 77.7) |
|  | Female | 20,450 | 16,101 | 78.7 (78.2 to 79.3) |
| School level | Kindergarten | 84 | 17 | 20.2 (13.0 to 30.0) |
|  | Primary | 11,933 | 10,283 | 86.2 (85.5 to 86.8) |
|  | Lower secondary | 17,854 | 14,023 | 78.5 (77.9 to 79.1) |
|  | Upper secondary | 9,826 | 6,613 | 67.3 (66.4 to 68.2) |
| Age group | 3 to 5 years | 84 | 17 | 20.2 (13.0 to 30.0) |
|  | 6 to 11 years | 16,608 | 14,222 | 85.6 (85.1 to 86.2) |
|  | 12 to 14 years | 13,179 | 10,084 | 76.5 (75.8 to 77.2) |
|  | 15 to 18 years | 9,826 | 6,613 | 67.3 (66.4 to 68.2) |
| Year | 2021 | 2,290 | 1,795 | 78.4 (76.7 to 80.0) |
|  | 2022 | 14,647 | 11,814 | 80.7 (80.0 to 81.3) |
|  | 2023 | 10,894 | 8,186 | 75.1 (74.3 to 75.9) |
|  | 2024 | 11,866 | 9,141 | 77.0 (76.3 to 77.8) |

Prevalence and 95% confidence intervals were calculated using the Wilson method.

**Figure 1.**
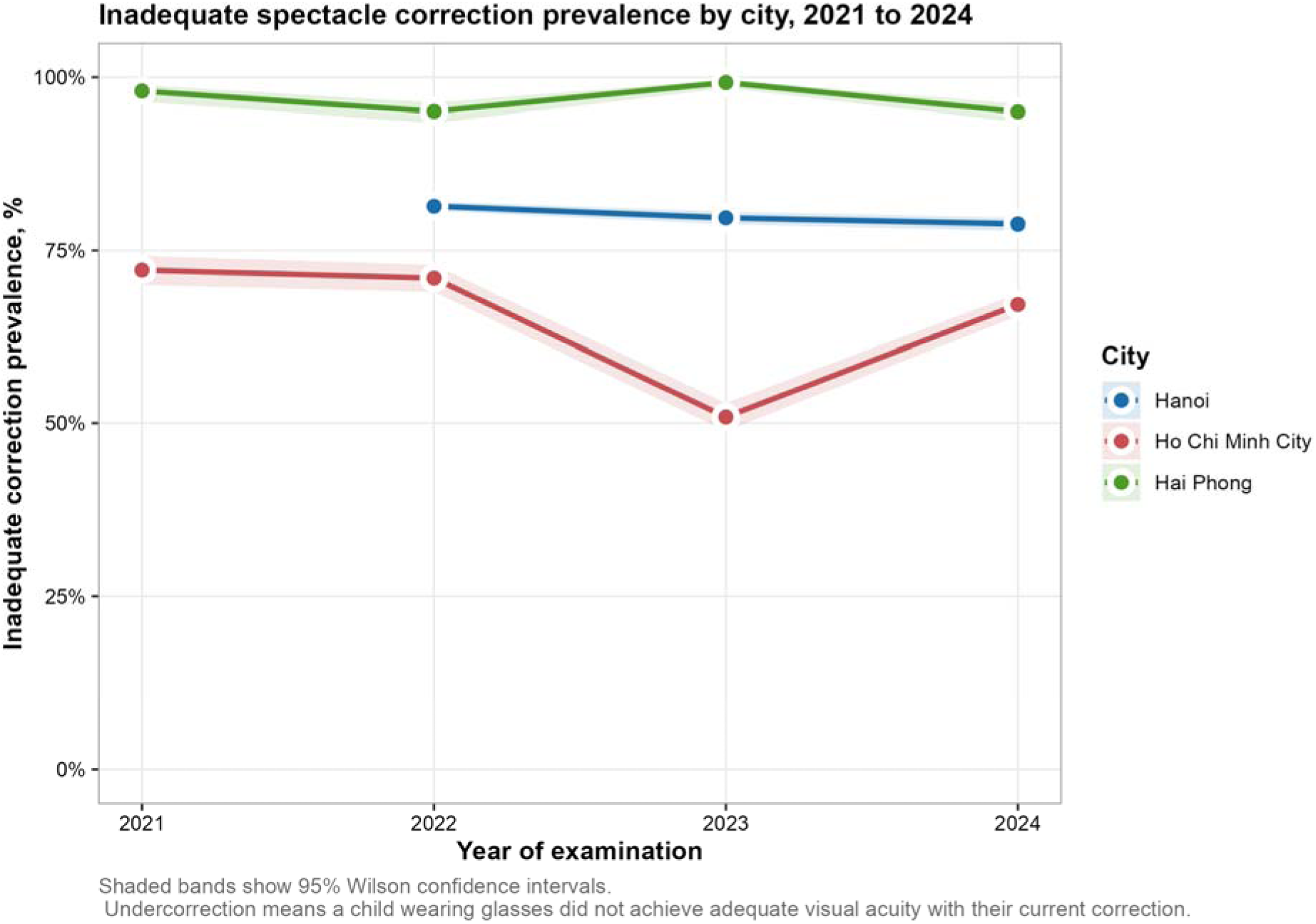
Inadequate spectacle correction prevalence by city, 2021 to 2024. Points show the annual prevalence of inadequate correction among spectacle-wearing children in each city, and shaded bands show 95% Wilson confidence intervals. Hanoi has no 2021 estimate because spectacle correction data collection began in that city in 2022. Inadequate correction means a child wearing glasses did not achieve adequate visual acuity with their current correction.

Inadequate correction was also strongly patterned by age. Prevalence peaked among primary school children aged 6 to 11 years at 85.6% (85.1 to 86.2), then declined steadily through lower secondary and upper secondary school to 67.3% (66.4 to 68.2) among the oldest children, aged 15 to 18 years (**Table 2**). The small kindergarten group showed a much lower prevalence of 20.2%, though this estimate rests on only 84 children and should be interpreted cautiously. The heat map in **Figure 2** makes this age gradient visible year by year, prevalence among 6 to 9 year olds sat consistently above 80% in most years, while prevalence among 16 to 18 year olds was consistently lower, in the 60s. Sex differences were modest, with inadequate correction slightly more common among girls than boys (78.7% versus 77.1%) (**Table 2**).

**Figure 2.**
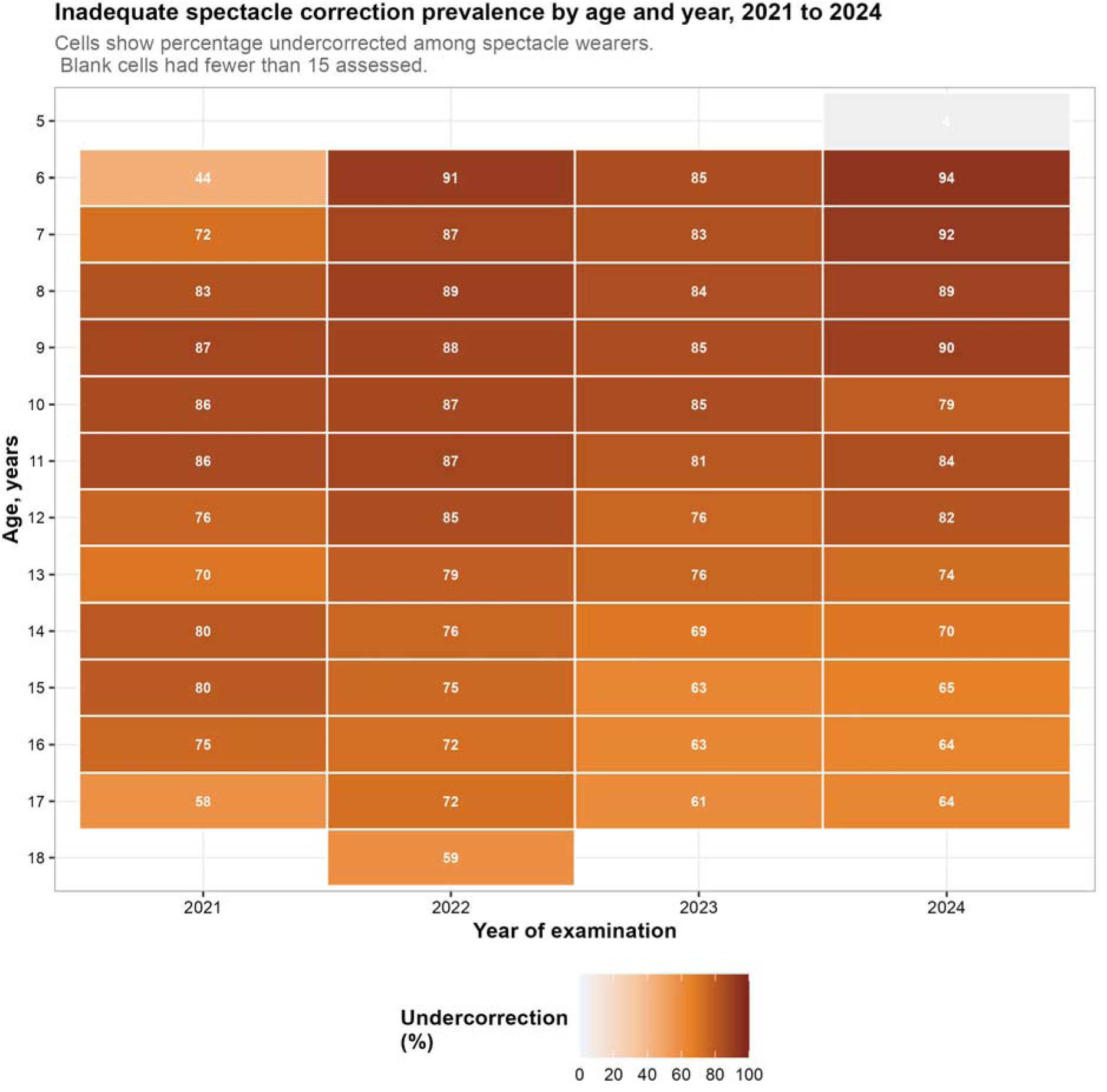
Inadequate spectacle correction prevalence by age and year, 2021 to 2024. Each cell shows the percentage of spectacle-wearing children at a given age and year who were inadequately corrected. Darker shading indicates higher prevalence. Blank cells indicate fewer than 15 children assessed at that age and year and were suppressed to avoid unstable estimates.

The primary adjusted model, a population-averaged modified-Poisson regression accounting for repeated examinations within the same child, confirmed these patterns after adjustment for calendar year, age, city, and sex (**Table 3**, **Figure 3**). Relative to Hanoi, the adjusted prevalence ratio for inadequate correction was 0.76 (95% CI= 0.75 to 0.78, p<0.001) in Ho Chi Minh City and 1.25 (95% CI= 1.23 to 1.26, p<0.001) in Hai Phong. Each additional year of age was associated with a small reduction in the adjusted prevalence ratio (aPR= 0.97 per year, 95% CI= 0.96 to 0.97, p<0.001), consistent with the crude age pattern. Girls remained at slightly higher adjusted risk than boys (aPR= 1.04, 95% CI= 1.03 to 1.06, p<0.001), and the prevalence of inadequate correction increased modestly across calendar years (aPR= 1.02 per year, 95% CI= 1.02 to 1.03, p<0.001).

**Table 3.** Factors associated with inadequate spectacle correction, child-level GEE.

| Term | aPR (95% CI) | p-value |
| --- | --- | --- |
| Calendar year (per 1-year increase) | 1.02 (1.02 to 1.03) | <0.001 |
| Age (per 1-year increase) | 0.97 (0.96 to 0.97) | <0.001 |
| City: Ho Chi Minh City | 0.76 (0.75 to 0.78) | <0.001 |
| City: Hai Phong | 1.25 (1.23 to 1.26) | <0.001 |
| Sex: Female | 1.04 (1.03 to 1.06) | <0.001 |

Primary population-averaged Generalized Estimating Equations (GEE) model among spectacle-wearing examinations. Outcome: inadequate correction (1 = yes). Reference categories: Hanoi and Male where both levels are present. Calendar year and age are continuous. Robust sandwich standard errors account for repeated examinations within child; working correlation = exchangeable. Adjusted prevalence ratios (aPRs) are reported.

**Figure 3.**
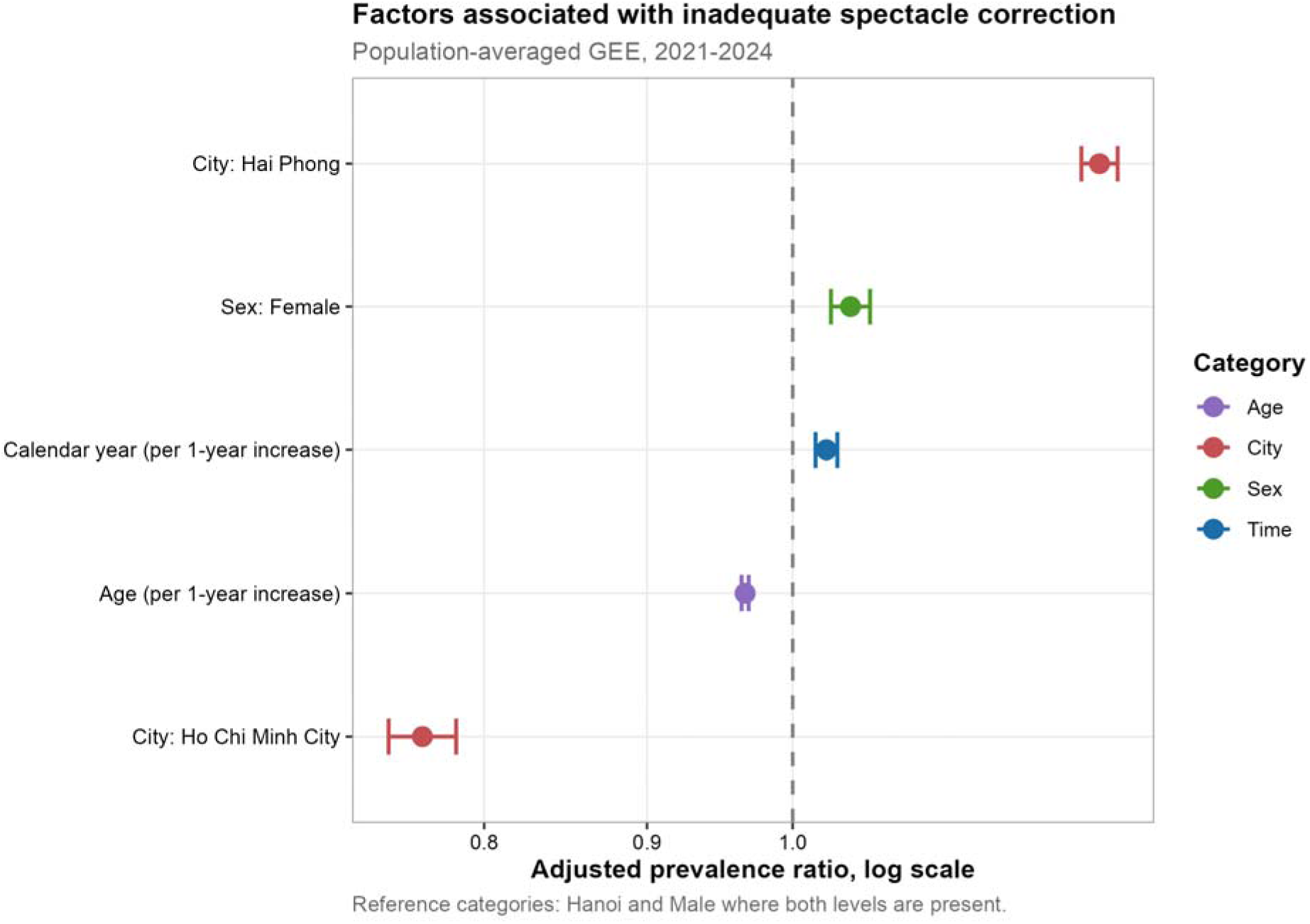
Factors associated with inadequate spectacle correction, population-averaged Generalized Estimating Equations (GEE), 2021 to 2024. Points show adjusted prevalence ratios and horizontal lines show 95% confidence intervals, plotted on a log scale. The dashed vertical line at 1.0 marks no association. Reference categories are Hanoi and Male.

The longitudinal linkage of 9,285 consecutive-year pairs from 8,341 children showed that inadequate correction, once present, tended to persist (**Table 4**, **Figure 4**). Among children who were inadequately corrected at their first visit, 85.3% (84.4 to 86.1) remained inadequately corrected the following year, and only 14.7% (13.9 to 15.6) achieved adequate correction. Children who started out adequately corrected had a less stable outlook than might be expected, with 53.4% (51.2 to 55.6) becoming inadequately corrected at their next visit. Among children who were inadequately corrected at baseline, the adjusted model in **Table 5** found that persistence was not related to calendar year (aPR= 0.99 per year, 95% CI= 0.98 to 1.00, p=0.187), but was somewhat lower in Ho Chi Minh City relative to Hanoi (aPR= 0.95, 95% CI= 0.92 to 0.99, p=0.006) and higher in Hai Phong (aPR 1.17, 1.15 to 1.20, p<0.001), broadly mirroring the pattern seen in the cross-sectional model.

**Table 4.** Annual transition in spectacle correction and probability of inadequate or adequate correction at the next annual visit.

| Status at first visit | Pairs | Probability inadequate next year, % (95% CI) | Probability adequate next year, % (95% CI) |
| --- | --- | --- | --- |
| Adequately corrected | 1,986 | 53.4 (51.2 to 55.6) | 46.6 (44.4 to 48.8) |
| Inadequately corrected | 7,299 | 85.3 (84.4 to 86.1) | 14.7 (13.9 to 15.6) |

Annual transition in spectacle correction status and probability of inadequate or adequate correction at the next annual visit among linked children. Pairs refers to the number of consecutive calendar-year observations contributing to each row. The clinically important quantities are the persistence of inadequate correction among children who started inadequately corrected, and the emergence of new inadequate correction among children who started adequately corrected.

**Table 5.** Factors associated with persistence of inadequate spectacle correction.

| Term | aPR (95% CI) | p-value |
| --- | --- | --- |
| Calendar year at first visit (per 1-year increase) | 0.99 (0.98 to 1.00) | 0.187 |
| Age (per 1-year increase) | 0.98 (0.98 to 0.99) | <0.001 |
| City: Ho Chi Minh City | 0.95 (0.92 to 0.99) | 0.006 |
| City: Hai Phong | 1.17 (1.15 to 1.20) | <0.001 |
| Sex: Female | 1.04 (1.02 to 1.06) | <0.001 |

Among children inadequately corrected at the first of two consecutive annual examinations. Outcome is inadequate correction at the next annual examination. Population-averaged Generalized Estimating Equations (GEE) accounts for repeated transitions within child.

**Figure 4.**
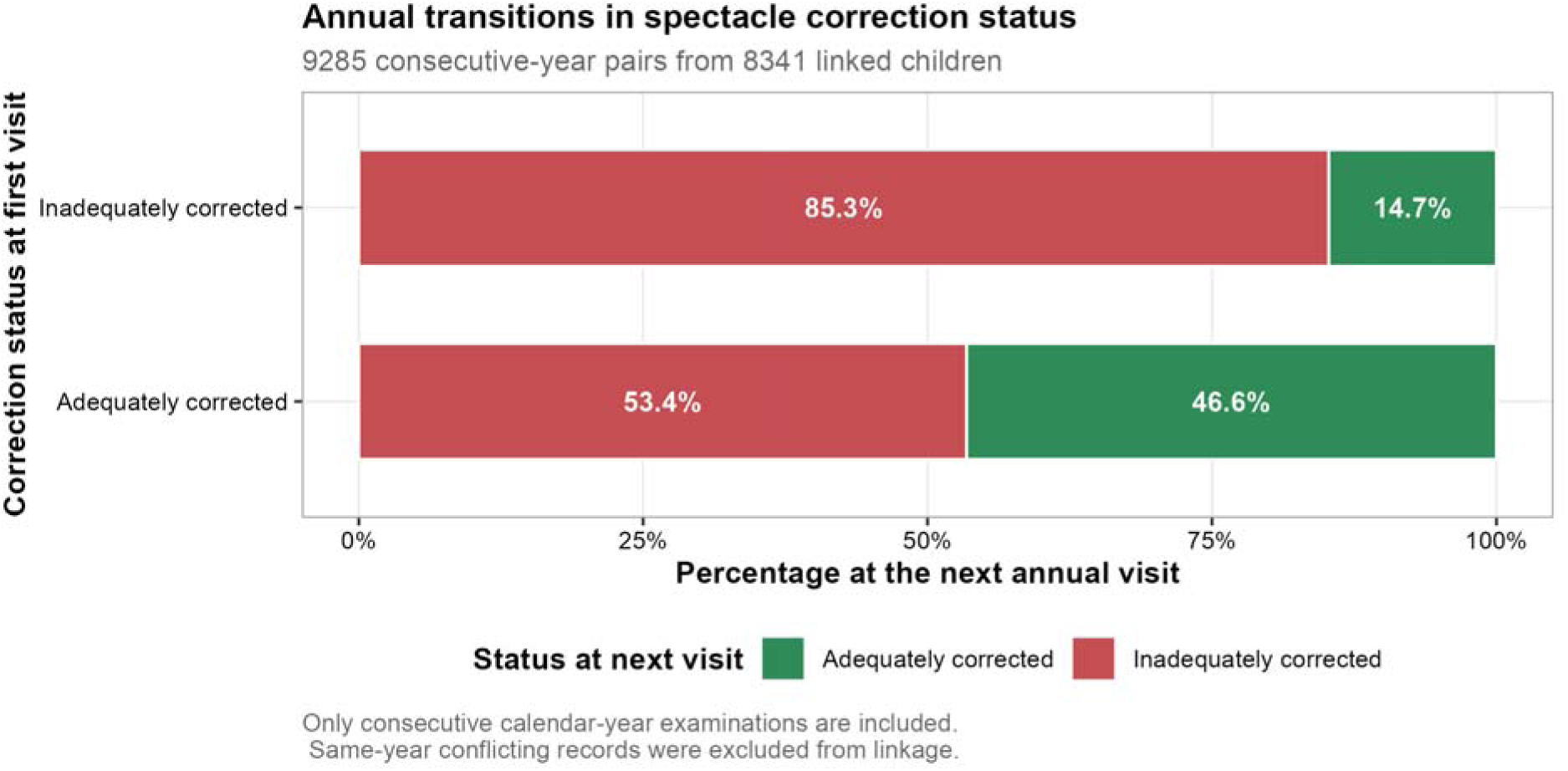
Annual transitions in spectacle correction status among linked children. Based on 9,285 consecutive-year pairs from 8,341 children. Bars show the percentage of children in each starting correction status who were adequately or inadequately corrected at their next annual visit. Only consecutive calendar-year examinations are included, and same-year records with conflicting correction status were excluded from linkage.

Sensitivity analyses supported the robustness of the main findings. A child-level mixed-effects logistic regression, which modeled a random intercept per child rather than the population-averaged approach used in the primary model, produced odds ratios pointing in the same direction for every covariate, including a markedly elevated odds ratio for Hai Phong (**Table S2**). Restricting the adjusted model to children aged 6 years and older left the estimated calendar-year effect essentially unchanged (aPR= 1.03 per year, 95% CI= 1.02 to 1.03) (**Table S3**), and the overall prevalence of inadequate correction in this restricted analysis, 77.4%, closely matched the 77.3% observed in the main analytic sample. **Table S1** shows that the proportion of examined children who wore spectacles ranged from about 24 to 38% across cities and years, with no obvious directional trend, suggesting that the patterns described above are unlikely to be explained by changes in who was captured in the spectacle-wearing denominator over time.

## Discussion

This multi-city analysis found that most Vietnamese schoolchildren who already wear glasses are still not adequately corrected, with prevalence ranging from about two thirds in Ho Chi Minh City to nearly universal inadequate correction in Hai Phong. These findings sit within a global pattern in which spectacles address the large majority of childhood vision impairment in principle, yet a high proportion of children who own glasses do not achieve adequate visual outcomes with them in practice ^2,3^. The scale of inadequate correction observed here is striking even against that backdrop, and it points to a gap between owning spectacles and being properly corrected that has received comparatively little attention next to the more commonly studied question of whether a child wears glasses at all.

The near tenfold city-level variation, with Hai Phong showing the highest burden and Ho Chi Minh City the lowest even after adjustment for age, sex, and calendar year, most likely reflects differences in local eye care infrastructure and the quality of refraction and dispensing services rather than differences in the children themselves. Cost, service quality, and the accessibility of follow-up refraction have long been identified as central barriers to effective correction in school-age populations ^1,10^. Compliance research has also emphasized that reasons for inadequate correction are highly context specific and shaped by local health system factors rather than by any single universal driver ^2^, which is consistent with the scale of variation found between three cities in the same country. Innovative delivery models built around social enterprise or vision center networks have shown promise in closing similar gaps elsewhere in low resource settings, and may be worth piloting in the cities where inadequate correction was most severe ^16^.

The age pattern observed, with inadequate correction concentrated among younger primary school children and gradually easing toward late adolescence, is plausible given how quickly refractive error can change during active growth, requiring more frequent prescription updates than many families are able to obtain. Existing literature on the relationship between age and spectacle compliance is inconsistent ^2^, and these results extend that literature by showing that even among children who do own glasses, adequacy of correction, not just willingness to wear them, follows a clear age gradient. The modest excess risk seen among girls in the adjusted model is worth noting alongside prior reports that girls tend to be more compliant wearers of spectacles once prescribed ^2,8^. Compliance and adequacy of correction are related but distinct constructs, and it is possible that girls in this cohort were more likely to wear their glasses consistently while still not having them updated as often as needed, though this study cannot distinguish between those explanations.

Perhaps the most concerning finding is that inadequate correction, once present, was highly persistent, with the large majority of affected children remaining inadequately corrected a year later. This matters because timely correction has been linked to meaningful gains in learning and wellbeing in systematic reviews ^9^ and in at least one Vietnamese cohort of adolescents ^11^, while the broader evidence base on school vision programs stresses that detection alone accomplishes little unless it is reliably connected to correction and follow-up ^3^. These data suggest that, in this setting, the connection between detection and adequate correction is not being made consistently for a large share of children.

These findings should be read alongside the near absence of Vietnam-specific evidence on this topic. To our knowledge, only one prior Vietnamese study has examined outcomes related to spectacle correction in children, and it focused on academic performance rather than correction adequacy itself ^11^. This study is limited by its reliance on a private school network, which may not represent the full range of eye care access across Vietnam, and by the absence of information on why individual children were inadequately corrected, whether due to non-wear, an outdated prescription, or a dispensing problem. Even so, given the strong economic case for investment in childhood eye health ^17^ and the WHO’s global push to expand effective refractive error coverage ^18^, these results argue for closer attention to correction quality, not just detection, in school eye health programs in Vietnam and similar countries ^3,19^.

## Data Availability

R code is available from the corresponding author upon reasonable request. Individual patient-level data cannot be shared due to applicable privacy regulations and the terms of the institutional ethics approval.

## DECLARATIONS

### Ethical Statement

The study was approved by the Vinmec Ethics Committee (approval number 0231/2024/CN/HDDD VMEC). Written informed consent was waived for secondary analysis of de-identified, retrospective data.

### Contributors

N.T.H. did conceptualization, data curation, formal analysis, investigation, methodology, project administration, resources, software, supervision, validation, visualization, writing original draft, and writing review & editing.

K.P.M.T. did data cleaning and writing review & editing.

All authors read and approved the manuscript.

### Use of Artificial Intelligence

All scientific content, analyses, and interpretations are the original work of the authors. The authors used AI-assisted tools for language editing and grammar checking during manuscript preparation.

## References

1. Sharma, A., Congdon, N., Patel, M. & Gilbert, C. School-based approaches to the correction of refractive error in children. Surv. Ophthalmol. 57 3, 272–283 (2012).

2. Morjaria, P., McCormick, I. & Gilbert, C. Compliance and Predictors of Spectacle Wear in Schoolchildren and Reasons for Non-Wear: A Review of the Literature. Ophthalmic Epidemiol. 26, 367–377 (2019).

3. Little, J.-A. et al. Current status of school vision screening—rationale, models, impact and challenges: a review. Br. J. Ophthalmol. 109, (2025).

4. Sankaridurg, P. et al. IMI Impact of Myopia. Invest. Ophthalmol. Vis. Sci. 62, (2021).

5. Morgan, I. et al. The epidemics of myopia: Aetiology and prevention. Prog. Retin. Eye Res. 62, 134 (2017).

6. Morgan, I. & Jan, C. China Turns to School Reform to Control the Myopia Epidemic: A Narrative Review. Asia-Pacific journal of ophthalmology (2022) doi:10.1097/apo.0000000000000489.

7. Evans, B., Shah, R. & Vlasak, N. The Changing Natural History of Anisometropia: A Scoping Review. Ophthalmic & Physiological Optics 46, 13–28 (2026).

8. Sheeladevi, S. et al. Prevalence of refractive errors in children in India: a systematic review. Clin. Exp. Optom. 101, (2018).

9. Pirindhavellie, G.-P., Yong, A., Mashige, K., Naidoo, K. & Chan, V. The impact of spectacle correction on the well-being of children with vision impairment due to uncorrected refractive error: a systematic review. BMC Public Health 23, (2023).

10. Burnett, A. et al. Interventions to improve school-based eye-care services in low- and middle-income countries: a systematic review. Bull. World Health Organ. 96, 682–694 (2018).

11. Elbasheer, A. D. H. E. et al. Refractive Errors and Educational Outcomes in Children: A Systematic Review. Cureus 18, (2026).

12. von Elm, E. et al. The Strengthening the Reporting of Observational Studies in Epidemiology (STROBE) statement: guidelines for reporting observational studies. J. Clin. Epidemiol. 61, (2008).

13. World Health Organization. Vision and Eye Screening Implementation Handbook. World Health Organization. Geneva: WHO Press (2024).

14. Liang, K. Y. & Zeger, S. L. Longitudinal data analysis using generalized linear models. Biometrika 73, (1986).

15. Zou, G. A Modified Poisson Regression Approach to Prospective Studies with Binary Data. Am. J. Epidemiol. 159, 702–706 (2004).

16. Morjaria, P. et al. Delivering Refractive Care to Populations With Near and Distance Vision Impairment: 2 Novel Social Enterprise Models. Asia-Pacific Journal of Ophthalmology 11, 59–65 (2022).

17. Wong, B. et al. The case for investment in eye health: systematic review and economic modelling analysis. Bull. World Health Organ. 101, 786–799 (2023).

18. Approaches for delivery of refractive and optical care services in community and primary care settings. Cochrane Database Syst. Rev. 2024, (2024).

19. Harvey, A.-A., Morjaria, P. & Tousignant, B. Priorities in school eye health in low and middle-income countries a scoping review. Eye 38, 1988–2002 (2024).

