## Supplementary Tables for "Prevalence, Predictors, and Persistence of Inadequate Spectacle Correction in Vietnamese Schoolchildren"

Nhan Thi Ho et al.

Table S1. Spectacle wearing and correction data availability by city and year

| City | Year | N total | N spectacle | % spectacle | N correction adequacy recorded |
| --- | --- | --- | --- | --- | --- |
| Hanoi | 2022 | 50,146 | 11,788 | 23.5 | 11,788 |
| Hanoi | 2023 | 28,970 | 7,635 | 26.4 | 7,635 |
| Hanoi | 2024 | 29,077 | 7,632 | 26.2 | 7,632 |
| Ho Chi Minh City | 2021 | 4,845 | 1,737 | 35.9 | 1,737 |
| Ho Chi Minh City | 2022 | 8,192 | 2,046 | 25.0 | 2,046 |
| Ho Chi Minh City | 2023 | 9,764 | 2,337 | 23.9 | 2,337 |
| Ho Chi Minh City | 2024 | 11,244 | 3,209 | 28.5 | 3,209 |
| Hai Phong | 2021 | 1,447 | 553 | 38.2 | 553 |
| Hai Phong | 2022 | 2,934 | 813 | 27.7 | 813 |
| Hai Phong | 2023 | 3,115 | 922 | 29.6 | 922 |
| Hai Phong | 2024 | 3,122 | 1,025 | 32.8 | 1,025 |

% spectacle is the percentage of all examined children in that city and year who were wearing spectacles at the time of examination.

Table S2. Child-level mixed-effects logistic regression sensitivity analysis

| Term | OR (95% CI) | p-value |
| --- | --- | --- |
| Year | 1.18 (1.11 to 1.26) | <0.001 |
| Age | 0.84 (0.83 to 0.85) | <0.001 |
| CityHo Chi Minh City | 0.38 (0.35 to 0.41) | <0.001 |
| CityHai Phong | 9.21 (7.37 to 11.51) | <0.001 |
| SexFemale | 1.24 (1.15 to 1.33) | <0.001 |

Random intercept for child identifier. This sensitivity model addresses within-child dependence through subject-specific random effects. It is secondary to the population-averaged Generalized Estimating Equations (GEE). OR= odds ratio, CI= confidence interval.

Table S3. Sensitivity analyses for inadequate spectacle correction

| Sensitivity check | N | Inadequate correction, % | Adjusted year aPR (95% CI) |
| --- | --- | --- | --- |
| Main analysis, confirmed RE spectacle wearers | 24,125 | 77.3 | 1.02 (1.02 to 1.03) |
| Age >=6 years, adjusted GEE | 24,059 | 77.4 | 1.03 (1.02 to 1.03) |

The primary model is the child-level Generalized Estimating Equations (GEE). Sensitivity analyses assess broader refractive error definition, quantitative-refraction restriction, and exclusion of children younger than 6 years. aPR= adjusted prevalence ratio, CI= confidence interval, RE= refractive error.
